# Low density neutrophils and neutrophil extracellular traps (NETs) are new inflammatory players in heart failure

**DOI:** 10.1101/2023.11.03.23298072

**Authors:** Benjamin L. Dumont, Paul-Eduard Neagoe, Elcha Charles, Louis Villeneuve, Sandro Ninni, Jean-Claude Tardif, Agnès Räkel, Michel White, Martin G. Sirois

## Abstract

**Background:** Heart failure with reduced (HFrEF) or preserved ejection fraction (HFpEF) is characterized by low-grade chronic inflammation. Circulating neutrophils regroup two subtypes termed high- and low-density neutrophils (HDNs and LDNs). LDNs represent less than 2% of total neutrophil under physiological conditions, but their count increase in multiple pathologies, releasing more inflammatory cytokines and neutrophil extracellular traps (NETs).

**Objectives:** Assess the differential count and role of HDNs, LDNs and NETs-related activities in HF patients.

**Methods:** HDNs and LDNs were isolated from human blood by density gradient and purified by FACS and their counts obtained by flow cytometry. NETs formation (NETosis) was quantified by confocal microscopy. Circulating inflammatory and NETosis biomarkers were measured by ELISA. Neutrophil adhesion onto human extracellular matrix (hECM) was assessed by optical microscopy.

**Results:** A total of 140 individuals were enrolled, including 33 healthy volunteers (HV), 41 HFrEF (19 stable patients and 22 presenting acute decompensated HF; ADHF) and 66 HFpEF patients (36 stable patients and 30 presenting HF decompensation). HDNs and LDNs counts were significantly increased up to 39% and 2740% respectively in HF patients compared to HV. In HF patients, the correlations between LDNs counts and circulating inflammatory (CRP, IL-6 and -8), Troponin T, NT-proBNP and NETosis components were all significant. In vitro, LDNs expressed more H3Cit and NETs and were more pro-adhesive, with ADHFpEF patients presenting the highest pro-inflammatory profile.

**Conclusions:** HFpEF patients present higher levels of circulating LDNs and NETs related activities, which are the highest in the context of acute HF decompensation.

**Clinical Perspective:**

- In comparison to HFrEF, HFpEF patients have higher levels of circulation LDNs and NETs-associated inflammatory cytokines, peaking in acute decompensated clinical condition.
- Furthermore, LDNs are producing more NETs and are more adhesive than HDNs, which can contribute to pro-thrombogenesis status described in HF patients
- Measurement of circulating NETs-associated biomarkers could become a novel tool to assess the the risk of acute thrombogenesis in hospitalized ADHFpEF patients.
- These measurements could lead to future clinical treatments using NETosis inhibitors alone or combined with NETs degradation enzymes (e.g. DNase I).
- At this time, additional preclinical studies are required to determine specific cell surface markers that could distinguish LDNs from HDNs in whole blood.
- Once available, circulating LDNs levels would be routinely measured and integrated in the complete blood count analysis to better assess patients’ inflammatory status.

## Introduction

Heart failure (HF) is a leading cause of morbidity and mortality worldwide. While existing treatments have notably enhanced survival rates and outcomes associated with HF and reduced ejection fraction (HFrEF), preventing major events in HF and preserved ejection fraction (HFpEF) remains an unmet clinical need^1–3^. Hence, a deeper understanding of HF-related mechanisms is important for therapeutic innovation.

Mounting evidence supports the role of inflammatory processes in HF^4, 5^. More specifically, recruited innate immune cells have been involved in myocardial remodeling and might increase risk for adverse clinical events among HF patients. Thus, the assessment of circulating leukocytes as a proxy of myocardial remodeling is becoming an emerging area of interest to predict clinical outcomes in HF patients^6, 7^.

Neutrophils are first-line responders of the innate immune system. Circulating neutrophils are classified in two subsets, the high-density neutrophils (HDNs) and the low-density neutrophils (LDNs)^8, 9^. LDNs normally represent less than 2% of neutrophils in healthy individuals^8, 9^, but their counts are increased in multiple pathological disorders^9, 10^. In addition, LDNs are more potent than HDNs to enhance inflammation by producing cytokines^10^ and neutrophil extracellular traps (NETs) formation (NETosis)^11, 12^. NETs are composed of double-stranded DNA decorated with pro-inflammatory cytokines and enzymes such as myeloperoxidase (MPO)^13^. Upon their release, NETs can bind to endothelial cells (ECs) through von Willebrand factor (vWF)^14, 15^ and P-selectin^16, 17^, providing a scaffold for the binding of platelets, neutrophils and erythrocytes, leading to fibrin deposition and thrombotic microvascular occlusion^12, 18^.

Elevated circulating neutrophils is associated with higher incidence of acute decompensated HF and mortality rate in HF patients^6, 7, 19^. The role of neutrophils in myocardial repair and its impact on patient’s prognosis have been reported in HFrEF patients^6, 20^. However, few data have been published in patients with HFpEF^7, 21^.

So far, these studies did not address the inflammatory role of LDNs in HF patients. Thus, our main objectives were to assess both circulating neutrophils subsets (LDNs and HDNs) and their NETs related activities in HF patients across the spectrum of heart failure with preserved and reduced ejection fraction.

## Methods

An expanded Methods section can be found in the Supplemental Appendix.

### Population

This was a prospective non-interventional study that included five groups of individuals: healthy volunteers (HV), stable heart failure (HF) patients with rEF or pEF (termed HFrEF and HFpEF), and acute decompensated heart failure (ADHF) patients with rEF or pEF (termed ADHFrEF and ADHFpEF). All participants were recruited at the MHI. This study was approved by the Scientific Research Committee and the Ethics Committee of the MHI (ethics No. ICM #01-406 and No. ICM #12-1374) and conforms to the principles outlined in the Declaration of Helsinki. Donors were informed about the procedures and signed a written informed consent before participating in the study.

### Selection criteria of healthy volunteers and patients

Healthy volunteers (HV) were included if they had no significant medical conditions and were not treated with anti-inflammatory or immunosuppressive drugs within 2 weeks before blood collection. The classification of phenotypes for the HF patients was in concordance with the latest European Society of Cardiology (ESC) guidelines^1^. HF patients were considered rEF or pEF if their left ventricular ejection fraction (LVEF) was ≤40% or ≥50% respectively, assessed by contrast ventriculography, magnetic resonance imaging, radionuclide ventriculography, or quantitative echocardiography within the previous 12 months assuming they presented no significant cardiac events since the last assessment. Stable HF patients with NYHA classification I to IV symptoms were recruited at the *Clinique d’Insuffisance Cardiaque* of the MHI, during their monitoring visit. Patients with ADHF were recruited via the emergency room or the medical wards at the MHI. The main inclusion criteria were as follow: 1) adult aged ≥18 years, 2) prior diagnosis of HFrEF or HFpEF (LVEF ≤40% or ≥50% respectively and evidence of structural heart disease), 3) admitted for ADHF with elevated NT-proBNP levels (N-terminal B-type natriuretic peptide ≥600 pg/mL (≥1800 pg/mL for patients with atrial fibrillation), 4) treated with at least one dose of intravenous diuretics.

The main exclusions criteria were a recent malignancy (<3 years) except basocellular carcinoma, acute inflammatory process such as an infection or episode of gout attack in the last 2 weeks, severe chronic pulmonary disease, severe renal failure (creatinine >250 µmol/L), liver dysfunction, a concomitant systemic inflammatory disease, anemia (hemoglobin <9 g/dL or hematocrit <25%), a recent cardiac surgery (<3 months), HF due to congenital malformation, a prospected lifespan <90 days, or any condition that might have prevented them from giving an informed consent.

### Serum and neutrophil collection

Venous blood from all participants was collected in serum separating tubes (SST) (3.5-ml blood volume) and in 30 mL syringes (containing 5 mL acid citrate dextrose for 25 mL whole blood). Additional details are available in the Supplemental Appendix. The SST tube was centrifuged to obtain serum, which was aliquoted and frozen at −80°C. Neutrophils were isolated using the Ficoll-Paque gradient method, as previously described^16^. Upon isolation, neutrophils were resuspended in phenol-free RPMI-1640 medium (Cambrex Bio Science, Walkersville, MD) supplemented with 25 mM HEPES (N-2-hydroxyethylpiperazine-N′-2-ethanesulfonic acid) (Sigma-Aldrich, Oakville, ON, Canada), 1% penicillin/streptomycin/ Glutamax (VWR Intl., Montreal, QC, Canada), 1 mM CaCl2 (BDH Chemicals, Toronto, ON, Canada) and 5% FBS (Fetal Bovine serum; VWR) (termed complete RPMI). Contamination by PMBCs was less than 0.1% as determined by morphological analysis and flow cytometry. Cell viability of neutrophils was greater than 98%, as assessed by Trypan blue dye exclusion assay.

### HDNs and LDNs isolation by cell sorting

The HDNs and PBMCs were resuspended at 10^7^ cells/mL in PBS and incubated for 20 min with the neutrophil marker (mouse anti-human CD66b) and a monocyte marker (mouse anti-human CD14) antibodies. Using a cell sorter (BD FACSARIA Fusion, BD Biosciences), LDNs (CD66b^+^ CD14^-/low^) were sorted. The purified HDNs and LDNs were counted by hemocytometer and resuspended at 10^6^ cells/mL in complete RPMI.

### NETosis assay by confocal microscopy

In 250 µL of complete RPMI medium, 50,000 HDNs or LDNs and agonists (PBS, PMA 25 nM and CRP 5 µg/mL) were incubated in 35mm Petri dishes for 60 min at 37°C with 5% CO_2_. A green nuclear fluorophore, non-permeable to live cells (SYTOX Green, 1 mM; Life Technologies, Burlington, ON, Canada), and a membrane coloring agent (WGA, Wheat Germ Agglutinin; 1 µg/mL, ThermoFisher) were added in 1X HBSS buffer as a dying solution. The supernatant of the Petri dishes was replaced by 250µL of dye-containing solution. The images were then captured by confocal microscopy (LSM 710; Carl Zeiss, Toronto, ON, Canada) and set to acquire a mosaic of pictures (5 × 5 images) (Zen 2; Carl Zeiss) (magnification, 200X). To quantify NETs area, the number of green pixels (i.e. NETs colored by Sytox green) and red pixels (e.g., surface covered by neutrophils, colored by WGA) were measured using Image Pro Premier 9.3 Software (Media Cybernetics, Rockville, MD, USA) with a threshold to exclude background fluorescence. The results were presented as a percentage of the surface covered by NETs relative to the surface covered by cells.

### Statistics

Group comparisons were evaluated using one-way ANOVA followed by a Dunnett post-hoc test on at least 3 independent experiments from independent donors. Alternatively, Brown-Forsythe correction followed by a Dunnett T3 post-hoc was used when applicable. The correlation between variables was analyzed using Pearson regression. Statistical significance was set at p = 0.05. Analysis was performed with GraphPad Prism 9 for Windows.

## Results

### Participant characteristics

A total of 140 patients were enrolled, including 33 healthy volunteers (HV), 42 HFrEF (19 stable patients and 22 patients presenting HF decompensation) and 66 HFpEF patients (36 stable patients and 30 presenting HF decompensation). The baseline characteristics are provided in Table 1. Most patients with stable HF exhibited NYHA class 2 symptoms while ADHF patients presented class 3 symptoms. There was no significant difference regarding NYHA class between HFrEF and HFpEF patients. Most HFrEF patients had underlying coronary artery disease. The proportion of patients presenting type 2 diabetes (DM) ranged from 43.3 to 63.2% among HF patients. NT-proBNP ranged from 2237 ± 376 ng/l in stable HFpEF group to 9671 ± 1785 ng/l in ADHFrEF group. NT-proBNP levels were significantly higher in patients with ADHF regardless of HF phenotype. Most patients were treated with anti-platelet and diuretic agents, statins. Angiotensin II receptor blockers (ARBs) were mostly used in HFrEF as opposed to HFpEF patients. Pearson correlation analyses showed no significant correlation between all circulating biomarkers, except for CRP and age in HV.

**Table I.** Clinical characteristics of the study population.

|  | HV<br>(n = 33) | stable HFrEF<br>(n = 19) | stable HFpEF<br>(n = 36) | ADHFrEF<br>(n = 22) | ADHFpEF<br>(n = 30) | p Value |
| --- | --- | --- | --- | --- | --- | --- |
| Age (years) | 37.30 ± 2.39 | 72.11 ± 1.92*** | 76.06 ± 1.38*** | 68.95 ± 3.04*** | 75.75 ± 1.97*** | <0.001 |
| Male | 17 (50%) | 13 (68.42%) | 19 (52.78%) | 14 (63.64%) | 16 (53.33%) | 0.6323 |
| NYHA class |  |  |  |  |  |  |
| II | - | 13 (68.42%) | 24 (66.67%) | 5 (22.73%)<sup>§</sup> | 4 (11.11%)<sup>§§</sup> | 0.087 |
| III | - | 4 (21.05%) | 12 (33.33%) | 12 (54.55%) | 21 (58.33%) | 0.1381 |
| IV | - | 0 (0%) | 0 (0%) | 4 (18.18%) | 3 (8.33%) | 0.047 |
| **Etiology** |  |  |  |  |  |  |
| Ischemia | - | 16 (84.21%) | 10 (27.78%)<sup>††</sup> | 11 (50%)<sup>§</sup> | 11 (30.56%)<sup>†</sup> | 0.0061 |
| Cardiomyopathy | - | 2 (10.53%) | 7 (19.44%) | 5 (22.73%) | 3 (8.33%) | 0.2816 |
| Valvular | - | 9 (47.37%) | 19 (52.78%) | 6 (27.27%) | 21 (58.33%)<sup>††</sup> | 0.0241 |
| Others | - | 9 (47.37%) | 12 (33.33%) | 11 (50%) | 19 (52.78%) | 0.5189 |
| LVEF | - | 32.27 ± 1.4 | 55.86 ± 0.83<sup>†††</sup> | 28.47 ± 2 | 55.24 ± 1.03<sup>†††</sup> | <0.001 |
| Hypertension (>130/70) | - | 7 (36.84%) | 14 (38.89%) | 14 (63.64%) | 14 (46.67%) | 0.2472 |
| Dyslipidemia | - | 15 (78.95%) | 21 (58.33%) | 14 (63.64%) | 25 (83.33%)<sup>§</sup> | 0.1101 |
| Diabete | - | 12 (63.16%) | 17 (47.22%) | 11 (50%) | 13 (43.33%) | 0.5826 |
| Overweight (BMI > 25) | - | 13 (68.42%) | 24 (66.67%) | 18 (81.82%) | 26 (86.67%) | 0.2076 |
| Obesity (BMI > 30) | - | 1 (5.26%) | 11 (30.56%)<sup>†</sup> | 11 (50%)<sup>§§</sup> | 15 (50%) | 0.0052 |
| **Hematology & Biochemistry** |  |  |  |  |  |  |
| Hemoglobin (g/L) | - | 132.67 ± 4.67 | 125.48 ± 2.38 | 125.95 ± 3.78 | 120.24 ± 3.2 | 0.1096 |
| Leukocytes (X10<sup>9</sup>/L) | - | 6.65 ± 0.49 | 6.81 ± 0.5 | 6.74 ± 0.51 | 8.63 ± 1.28 | 0.222 |
| CBC neutrophils (X10<sup>9</sup>/L) | 3.41 ± 0.18 | 4.26 ± 0.40 | 4.77 ± 0.42* | 5.03 ± 0.45** | 6.20 ± 0.91** | 0.0086 |
| Lymphocytes (X10<sup>9</sup>/L) | 2.12 ± 0.27 | 1.19 ± 0.12** | 1.05 ± 0.10*** | 0.96 ± 0.08 *** | 1.26 ± 0.14** | <0.001 |
| CBC neutrophils/lymphocytes ratio | 2.04 ± 0.18 | 3.61 ± 0.78** | 4.54 ± 0.62*** | 5.24 ± 0.83 *** | 4.84 ± 0.49*** | <0.001 |
| K<sup>+</sup> (mmol/L) | - | 4.46 ± 0.14 | 4.25 ± 0.07 | 3.95 ± 0.09<sup>§§</sup> | 4.17 ± 0.11 | 0.0179 |
| Na<sup>+</sup> (mmol/L) | - | 138.56 ± 0.75 | 136.51 ± 0.73 | 138.4 ± 0.86 | 137.49 ± 0.57 | 0.1687 |
| Cl<sup>-</sup> (mmol/L) | - | 106.06 ± 0.9 | 102.37 ± 0.91<sup>†</sup> | 103 ± 1.07 | 101.59 ± 0.84 | 0.0181 |
| Urea (mmol/L) | - | 13.15 ± 1.49 | 15.17 ± 1.43 | 16.81 ± 1.96 | 14.76 ± 1.35 | 0.5443 |
| Creatinine (μmol/L) | - | 152.83 ± 9.91 | 141.54 ± 8.24 | 146.45 ± 11.72 | 132.45 ± 8.6 | 0.5237 |
| NT-proBNP (ng/L) | 32.1 ± 4.6 | 3406 ± 737*** | 2237 ± 376*** | 9671 ± 1785***/<sup>§§§</sup> | 5873 ± 1334 ***/<sup>§§</sup> | < 0.001 |
| Troponin T (ng/L) | 5.6 ± 1.16 | 34.7 ± 6.07*** | 26.8 ± 2.75*** | 49.7 ± 4.86*** | 48.6 ± 6.33*** | < 0.001 |
| G-CSF(ng/L) | 65.8 ± 3.6 | 80.9 ± 9.3 | 68.3 ± 3.8 | 79.9 ± 4.4 | 69.8 ± 2.5 | 0.156 |
| **Medication** |  |  |  |  |  |  |
| ACEi | - | 0 (0%) | 2 (5.56%) | 1 (4.45%) | 6 (20%) | 0.0507 |
| ARB | - | 15 (78.95%) | 9 (25%)<sup>†††</sup> | 9 (40.91%)<sup>§</sup> | 2 (6.67%)<sup>††</sup> | < 0.001 |
| Beta-blocker | - | 19 (100%) | 25 (69.44%)<sup>†</sup> | 19 (86.36%) | 26 (86.67%) | 0.0278 |
| Diuretic agent | - | 15 (78.95%) | 32 (88.89%) | 18 (81.82%) | 25 (83.33%) | 0.7785 |
| Statin | - | 16 (84.21%) | 19 (52.78%)<sup>†</sup> | 13 (59.09%) | 23 (76.67%) | 0.0542 |
| Anti-platelet & Anti-coagulant | - | 15 (78.95%) | 32 (88.89%) | 14 (63.64%) | 22 (73.33%) | 0.1425 |
| Metformin | - | 4 (21.05%) | 7 (19.44%) | 1 (4.45%) | 7 (23.33%) | 0.3234 |
| Sulfonylurea | - | 3 (15.79%) | 3 (8.33%) | 0 (0%) | 2 (6.67%) | 0.2907 |
| α-glucosidase inhibitor | - | 0 (0%) | 0 (0%) | 0 (0%) | 0 (0%) | 0.9999 |
| DPP-4 inhibitor | - | 5 (26.32%) | 8 (22.22%) | 3 (13.64%) | 8 (26.67%) | 0.7767 |
| GLP-1 agonist | - | 0 (0%) | 0 (0%) | 0 (0%) | 0 (0%) | 0.9999 |
| SGLT2 inhibitor | - | 5 (26.32%) | 4 (11.11%) | 2 (9.09%) | 0 (0%) | 0.0321 |
| Insulin | - | 3 (15.79%) | 6 (16.67%) | 7 (31.82%) | 6 (20%) | 0.4488 |

HF vs CTL : \* p < 0.05, \*\* p < 0.01, \*\*\*p < 0.001; HFpEF vs HFrEF : † p < 0.05; †† p < 0.01; ††† p < 0.001; ADHF vs stable HF : § p < 0.05, §§ p < 0.01, §§§ p < 0.001.

### Circulating counts of LDNs and HDNs and lymphocytes

From the complete blood count (CBC) measurements (Table 1), the total count of neutrophils in HV (3.41 x 10^9^/L) was significantly higher in HFpEF, ADHFrEF and ADHFpEF patients (4.26 – 6.20 x 10^9^/L). The total count of lymphocytes in HV (2.06 x 10^9^/L) was significantly lower in all HF cohorts (0.96 - 1.26 x 10^9^/L), and the ratio of neutrophils over lymphocytes (N/L) ranged from 2.04 (HV) to 3.61 - 5.24 in HF patients (p<0.01) (Table 1). Using flow cytometry, we assessed the total number of neutrophils (HDNs + LDNs), HDNs and LDNs in all groups (Figure 1). Compared to HV, the total count of neutrophils and HDNs was higher in all HF groups (significantly in HFpEF, ADHFrEF and ADHFpEF) (total neutrophils p<0.001) (Figure 1A) and up to 1.39-fold higher (HDNs p<0.05) in ADHFpEF patients (Figure 1B). The count of LDNs was significantly higher in all HF cohorts, up to 27-fold higher in ADHFpEF (p<0.01) (Figure 1C). The HDNs levels were similar among HF cohorts, whereas LDNs count in HFpEF were 2-fold higher compared to HFrEF and by 1.15-fold higher in ADHFpEF compared to ADHFrEF (Figure 1C).

**Figure 1.**
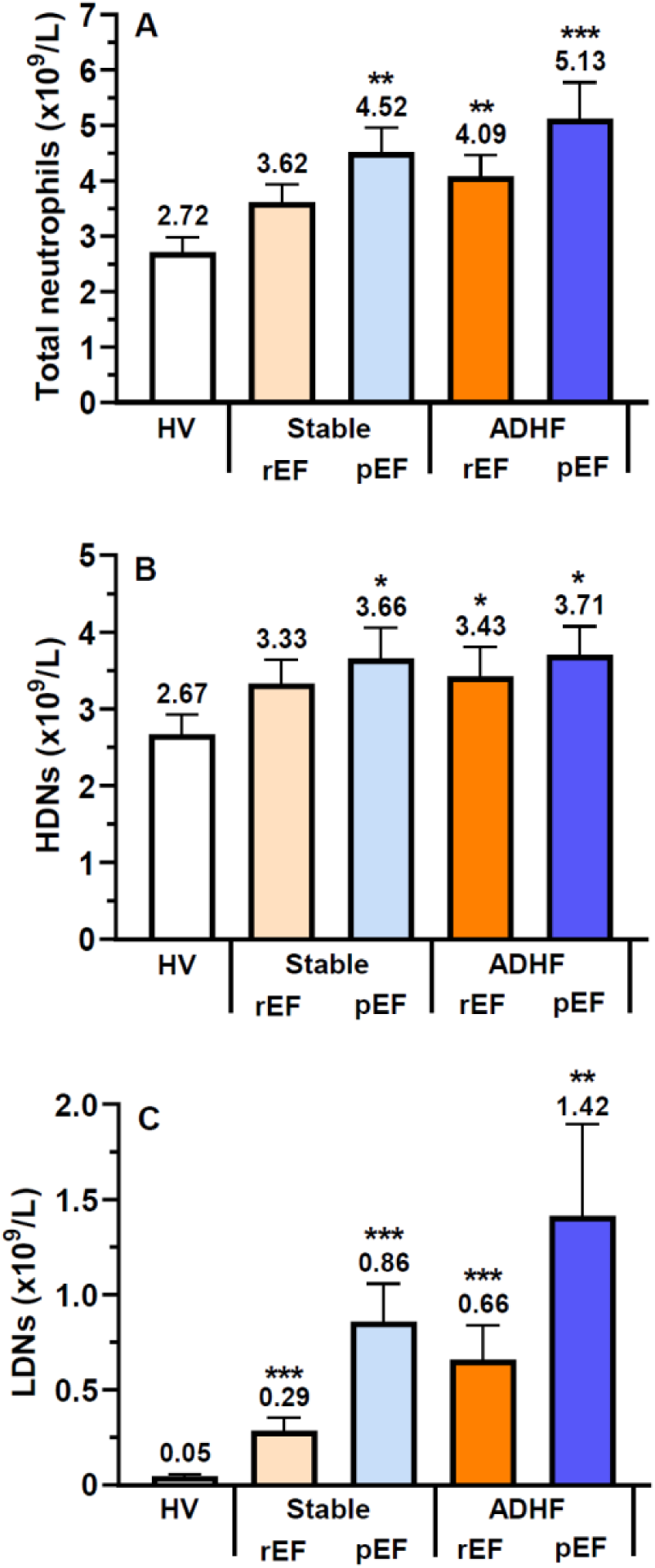
Circulating neutrophils, HDNs, LDNs and their respective lymphocytes ratio. Isolated (A) total neutrophils, (B) HDNs and (C) LDNs counts were determined by flow cytometry. The data are expressed as the absolute number of cells per liter of blood. All values are presented as mean ± SEM. P<0.05 was considered statistically significant (∗P<0.05, ∗∗P<0.01, ∗∗∗P<0.001 vs HV).

### Circulating biomarkers of inflammation and NETs

Circulating granulocyte colony stimulating (G-CSF) was quantified to ensure that neutrophil increase (neutrophilia) observed in HF patients was not due to infection-associated inflammation^22–24^. G-CSF levels did not change significantly between HV and all HF subgroups (Table 1). Circulating NETs were measured using H3Cit and MPO-DNA, two specific NETs components and MPO, an enzyme involved NETs formation^16, 17, 25^. All three biomarkers were significantly increased in all HF groups with maximal values observed in ADHFpEF patients. These NETosis biomarkers were significantly higher in ADHFpEF compared to HFpEF patients (Figure 2A-C). In all HF groups, we observed a significant increase of circulating IL-6, IL-8 and CRP known for their capacity to induce NETosis^13, 26, 27^ with maximal values in ADHFpEF patients (Figure 2D-F). In ADHFrEF the levels of IL-6 and IL-8 were significantly higher compared to HFrEF patients, whereas those markers were all significantly increased in ADHFpEF compared to HFpEF patients (Figure 2A-F).

**Figure 2.**
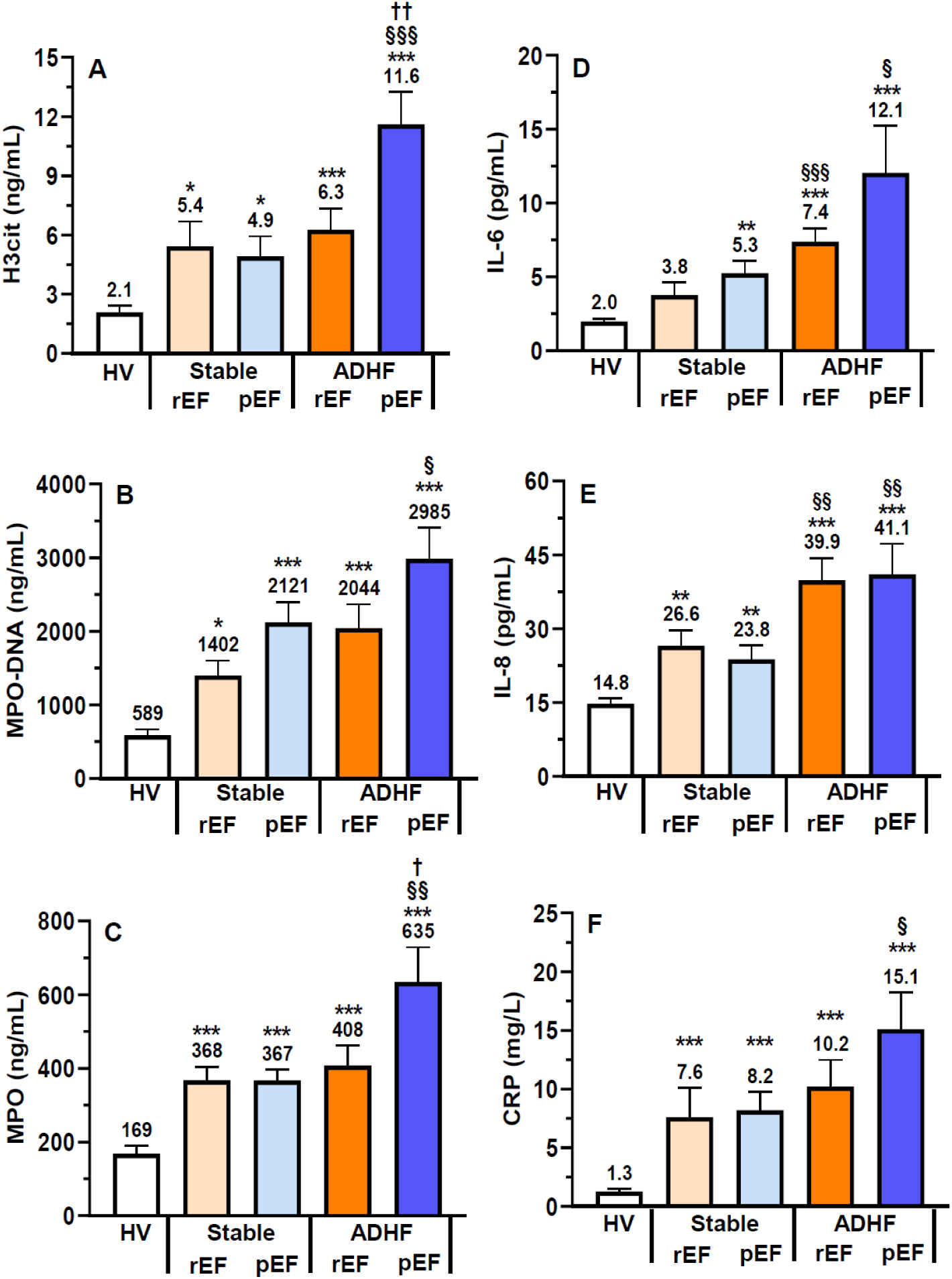
Circulating NETs-associated biomarkers and NETs-inducing cytokines. Circulating NETs-associated biomarkers (A) Citrullinated histone H3 (H3Cit), (B) myeloperoxidase-DNA complex (MPO-DNA) and C) MPO were measured in serum by ELISA. Circulating NETs-inducing cytokines (D) interleukin-6 (IL-6) and IL-8 were measured by a multiplex assay, whereas (F) C-reactive protein (CRP) was quantified by nephelometry. All values are presented as mean ± SEM. P<0.05 was considered statistically significant (∗P<0.05, ∗∗P<0.01, ∗∗∗P<0.001 vs HV; §P<0.05, §§P<0.01, §§§P<0.001 vs stable counterpart; †P<0.05, ††P<0.01 vs ADHFrEF).

Using a heat map, we represented the correlation between total neutrophils, HDNs, LDNs and circulating biomarkers. Both total neutrophils and LDNs strongly correlated with all biomarkers in a significant manner, except for G-CSF (not significant), whereas HDNs presented a weaker significant correlation with all biomarkers, except for G-CSF, IL-8 and NT-proBNP (not significant) (Supplemental Figure 1).

### Basal NETosis in isolated HDNs and LDNs

We assessed the percentage of HDNs and LDNs undergoing basal NETosis (HDNs-H3Cit^+^ and LDNs-H3Cit^+^) (Figure 3A), and their corresponding levels of expression measured by H3Cit^+^ mean fluorescence intensity (MFI) (Figure 3B). We observed a higher percentage of HDNs-H3Cit^+^ and LDNs-H3Cit^+^ in all 4 HF subgroups compared to HV, and the percentage of LDNs-H3Cit^+^ was higher than HDNs-H3Cit^+^ in HV and HF groups (Figure 3A). The H3Cit^+^ MFI of LDNs from all HF groups was significantly higher (∼4 to 11-fold) compared to their corresponding HDNs. Furthermore, H3Cit^+^ MFI of LDNs from HF patients were also up to 4-fold higher than HV (Figure 3B).

**Figure 3.**
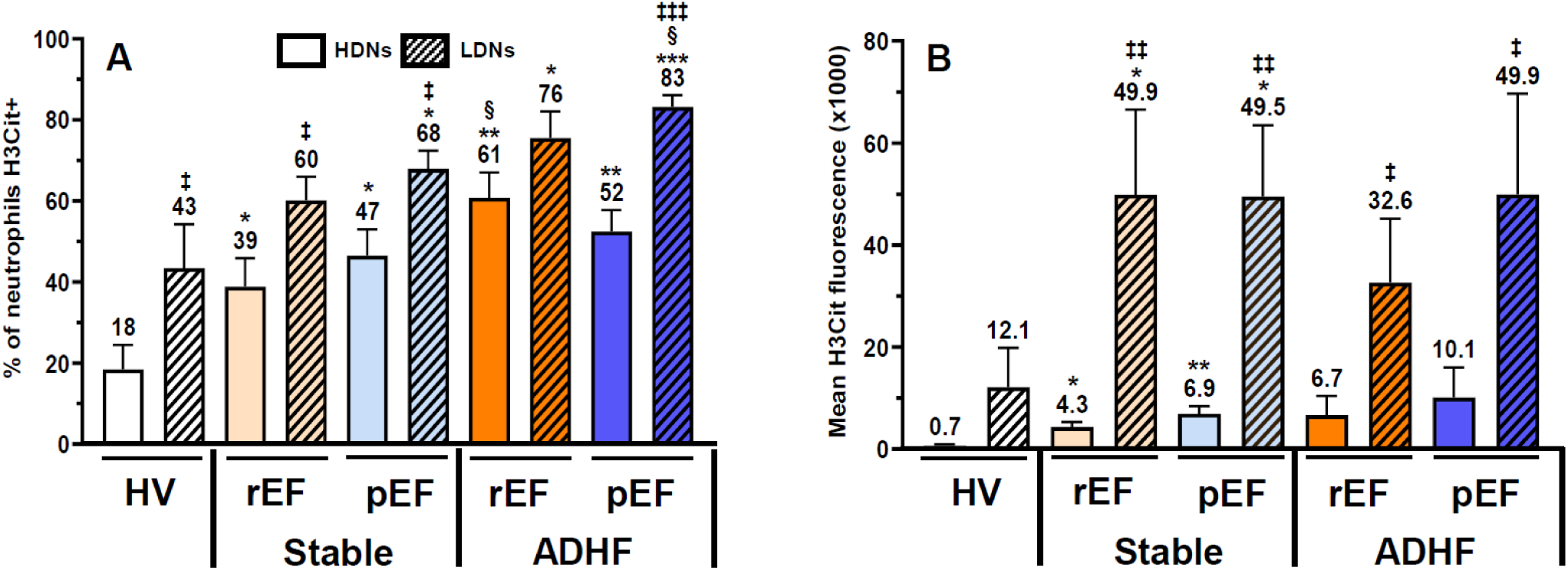
Basal NETosis in isolated HDNs and LDNs. (A) Isolated HDNs and LDNs were incubated with a NETs-associated biomarker (anti-H3Cit Ab) and flow cytometry was used to determine the percentage of cells in NETosis. The data are expressed as the percentage of H3Cit positive cells. (B) Using the same data collected by flow cytometry, the fluorescence intensity of H3Cit was measured to determine the relative expression of NETs at the cell surface. The data are expressed as fluorescence intensity (FI). All values are presented as mean ± SEM. P<0.05 was considered statistically significant (∗P<0.05, ∗∗P<0.01, ∗∗∗P<0.001 vs HV; ‡P<0.05 and ‡‡‡P<0.001 vs HDNs; §P<0.05 vs stable HF counterpart).

### In vitro NETosis from stimulated HDNs and LDNs

Neutrophils (HDNs and LDNs) were exposed to PBS (control vehicle) and pro-inflammatory agonists (PMA; 25 nM and CRP; 5 mg/mL) for 60 min and quantified by confocal microscopy (representative images Figure 4A and supplemental Figure 2). Under PBS stimulation, HDNs and LDNs from all HF groups significantly overexpressed NETs compared to corresponding HDNs and LDNs from HV (Figure 4B). In addition, NETosis in LDNs from ADHFrEF and ADHFpEF was significantly higher compared to their corresponding HDNs. Finally, NETosis in LDNs from ADHFpEF was significantly higher compared to LDNs from HFpEF (Figure 4B). Stimulation with PMA increased significantly NETosis in HDNs (except for ADHFrEF) and LDNs from all HF patients, compared to PBS. In all HF patients, there was a higher PMA-mediated NETosis in LDNs compared to HDNs counterparts (Figure 4C-D). Compared to PBS, CRP was more efficient to promote NETosis in LDNs vs HDNs from all HF patients (significant increase in HFrEF and HFpEF, and ADHFpEF) (Figure 4C-D).

**Figure 4.**
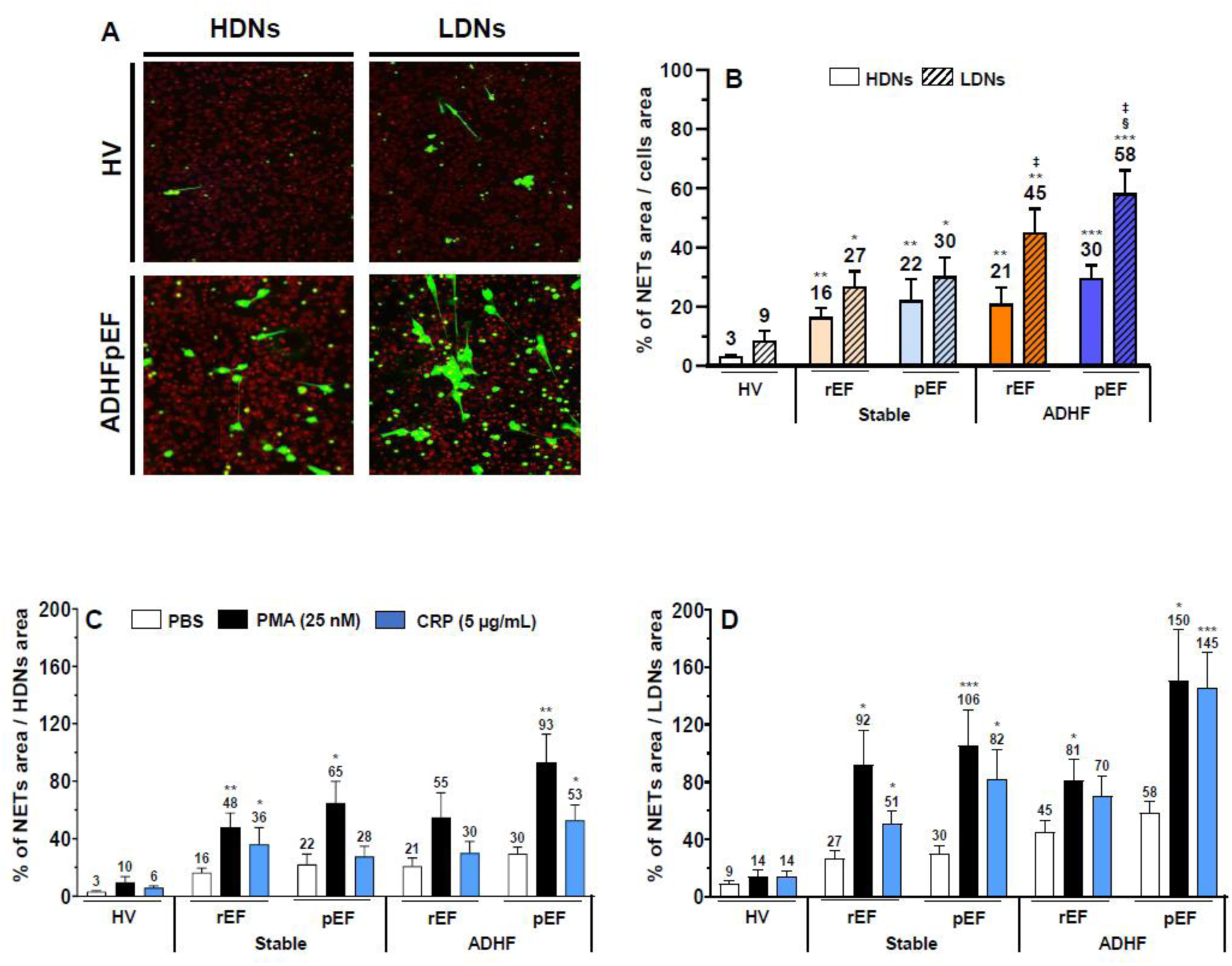
*In-vitro* NETosis from isolated HDNs and LDNs. Isolated HDNs and HDNs were treated with PBS, PMA and CRP for 1 hour, then stained with Sytox Green for DNA staining and WGA for cell membrane staining. The images were then captured by confocal microscopy and set to acquire a mosaic of pictures (5 × 5 images at 200X magnification). (A) Representative images of HDNs, LDNs (stained with WGA – red color) and NETs (stained with Sytox Green – green color). NETs were quantified by counting the number of green pixels (Sytox Green) and red pixels (WGA) using Image Pro Premier 9.3 Software with a threshold to exclude background fluorescence. (B) HDNs and LDNs basal (PBS), (C) HDNs and (D) LDNs agonist-induced NETosis was expressed as a percentage of the surface covered by NETs relative to the surface covered by cells. All values are presented as mean ± SEM. P<0.05 was considered statistically significant (∗P<0.05, ∗∗P<0.01, ∗∗∗P<0.001 vs HV-corresponding agonist; §P<0.05 vs stable HF counterpart; ‡P<0.05 vs HDNs).

### Binding of HDNs and LDNs onto human extracellular matrix (hECM)

HDNs and LDNs were exposed to PBS, PMA (25 nM) or CRP (5 mg/mL), transferred onto hECM-coated plates and incubated for 7.5 min. Under PBS stimulation, adhesion of HDNs and LDNs from all HF groups was significantly increased, up to 4.5- and 8-fold respectively in ADHFpEF compared to HV. HDNs and LDNs from ADHFpEF were significantly more adhesive compared to HFrEF, HFpEF and ADHFrEF (Figure 5A). HDNs and LDNs isolated from ADHFpEF were the most adherent upon stimulation with PMA or CRP (Figure 5B-E; Supplemental Figure 3). PMA is known to promote the activation of neutrophil β_2_-integrin (CD11b/CD18) complex, whereas CRP induces NETosis, therefore, we assessed if a pretreatment with a blocking goat anti-human CD18 Ab would reduce both HDNs and LDNs adhesion onto hECM. NETs were also degraded with DNase I^16^ to assess HDNs and LDNs cell-surface NETs contribution to hECM adhesion. Anti-CD18 and DNase I combination was used to assess their dual capacity to prevent HDNs and LDNs adhesiveness. Only DNase I combined with anti-CD18 significantly reduced basal adhesion of HDNs from ADHFpEF onto hECM by up to 39% (Figure 5B-E). In LDNs, all treatments (anti-CD18 and DNase I, alone or combined) significantly reduced basal adhesion up to 59% (Figure 5E). In addition, the combination of anti-CD18 and DNase I was significantly more effective to prevent LDNs adhesion compared to each pretreatment alone (Figure 5E). In HV, following PMA stimulation, only the combination of anti-CD18 and DNase I significantly reduced both HDNs and LDNs adhesion by 58 and 73% respectively (Figure 5B-C). Following CRP stimulation, only the combination of anti-CD18 and DNase I significantly reduced LDNs adhesion by 75% (Figure 5C). In HDNs and LDNs from ADHFpEF, all treatments (anti-CD18 and DNase I, alone or combined) significantly reduced PMA- or CRP-induced adhesion from 56 to 70% (Figure 5D-E). Again, dual pretreatment (anti-CD18 and DNase I) was significantly more effective to prevent HDNs and LDNs PMA- or CRP-induced adhesion compared to each pretreatment alone (Figure 5D-E). The adhesion profile of HDNs and LDNs from HFrEF and HFpEF and ADHFrEF patients are summarized in Supplemental Figure 3.

**Figure 5.**
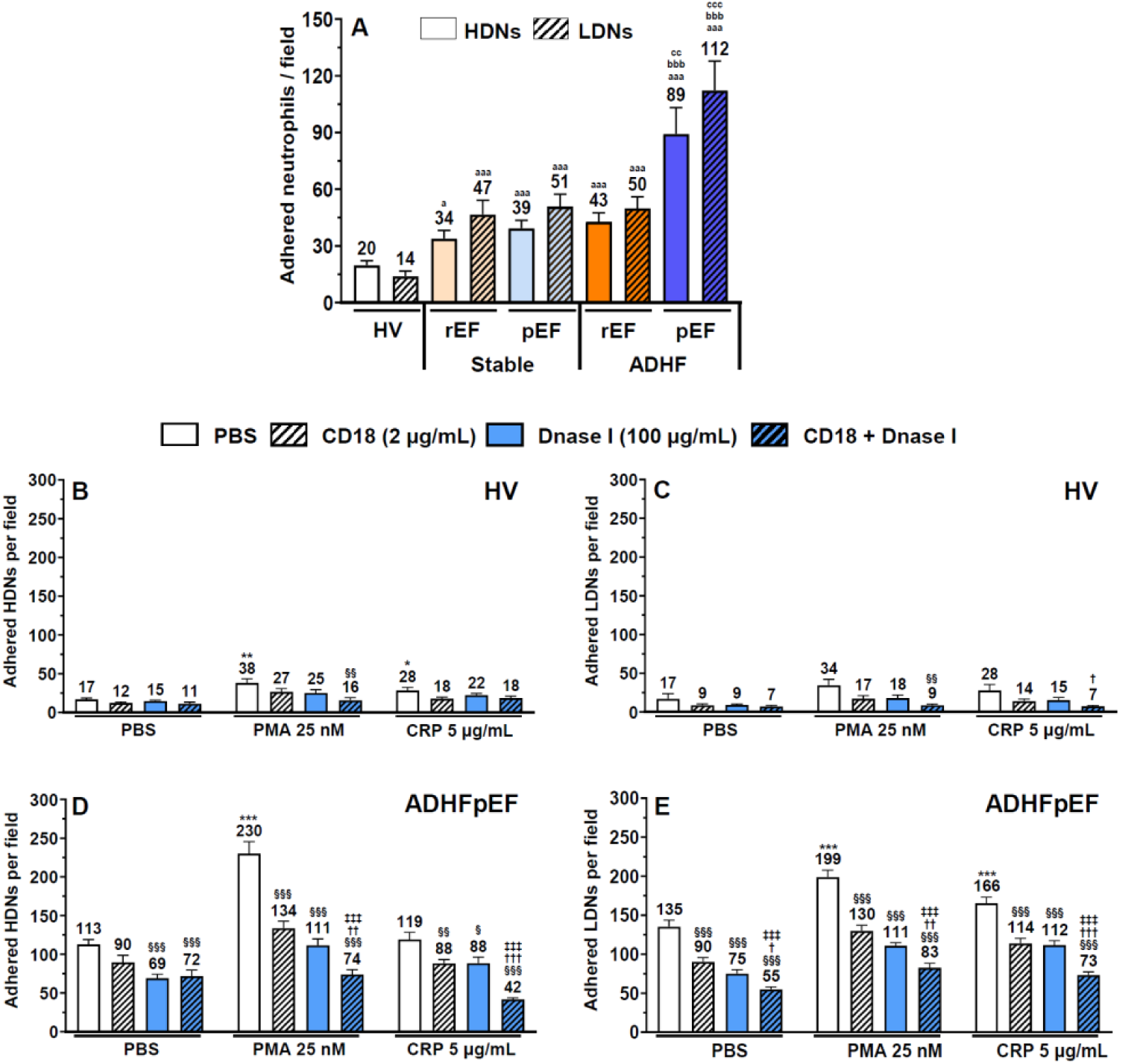
*In-vitro* isolated HDNs and LDNs adhesion to hECM. Isolated HDNs and HDNs were pretreated with anti-human ß2 integrin/CD18 Ab and/or DNase I for 30 minutes prior to incubation with PBS, PMA and CRP for an additional 7.5 minutes on hECM coated 48-plates. Adhered neutrophils were counted in 4 fields of view per well by optical microscopy using a digital camera. Adhesion of (A) basal (PBS) HDNs and LDNs agonist-induced, (B) HDNs and (C) LDNs from HV and (D) HDNs and (E) LDNs from ADHFpEF was expressed by the average number of adhered neutrophils / field from the four FOV of each well. All values are presented as mean ± SEM. P<0.05 was considered statistically significant. (aP<0.05, aaaP<0.001 vs corresponding HV; bbbP<0.001 vs stable HF counterpart; ccP<0.01, cccP<0.001 vs ADHFrEF; ∗P<0.05, ∗∗P<0.01, ∗∗∗P<0.001 vs PBS; §P<0.05, §§P<0.01, §§§P<0.001 vs PBS pre-treatment; †P<0.05, ††P<0.01, †††P<0.001 vs DNase I; ‡‡‡P<0.001 vs CD18).

## Discussion

Our study is the first to investigate the differential levels of HDNs and LDNs in HF patients. All HF patients presented high levels of circulating LDNs (up to 28-fold compared to HV), whereas the level of HDNs were 1.4-fold higher than HDNs levels in HV. The correlations between LDNs counts, circulating inflammatory cytokines, NETs components and their adhesiveness to hECM were all significantly increased in HF patients compared to HV. Patients presenting decompensated HF presented higher levels of these components, being the highest in ADHFpEF patients.

Chronic inflammation is common in HF. Changes in white blood cells count and their subtypes, such as neutrophilia are associated with increased risk of cardiovascular events and correlates with higher incidence of ADHF and mortality rate in HF patients^6, 7, 19^. LDNs have been previously linked to various pro-inflammatory disorders. For example, increased LDNs and NETs levels are found in patients with lupus erythematosus and might be involved in vascular damage and atheroma plaques rupture in this context^28, 29^. In cancer patients, elevated levels of circulating NETs positively correlate with interleukins (IL)-6 and -8 and a 2-fold increased risk of short-term mortality^30^. A positive correlation between circulating NETs and both intensive care unit hospitalization and mortality was also found in COVID-19 patients^31, 32^.

In agreement with previous studies, we observed little to none circulating LDNs in healthy volunteers^8, 9^, whereas LDNs counts were markedly increased in all HF patients. In addition, circulating LDNs were notably increased in acute decompensated HF, irrespective of LVEF status, contrasting with slight changes in HDNs counts. Interestingly, LDN subsets exhibited a significant rise in NETs-related activity in comparison to HDNs, a difference that was further heightened in patients experiencing decompensated HF.

NETs synthesis was previously associated with various thrombotic processes^18, 33^. In a recent study, we found that patients undergoing lung transplants with stage 3 primary graft dysfunction (PGD3) had increased NETosis biomarkers (MPO-DNA, MPO, and cfDNA) and pro-inflammatory cytokines (IL-8 and IL-6) correlating with severe graft dysfunction. In addition, as depicted by immunofluorescence staining, we observed in some of the lung graft tissues prior to transplantation, microvascular occlusion mainly derived from neutrophils undergoing NETosis and platelet-bound neutrophils adhered onto lung endothelium^34^.

Considering that NETs formation is involved in such thrombotic processes and given that HF decompensation is a high-risk situation for thromboembolic events, one could hypothesize the potential involvement of LDNs in the thrombotic risk associated with acute decompensated HF. This raises the possibility of LDNs serving as a valuable factor in assessing thrombotic risk among patients with HF.

We previously reported a marked increase of spontaneous NETosis in neutrophils isolated from patients with HF and found a positive correlation between circulating IL-6, IL-8, CRP and NETs levels. However, only NETs synthesis from isolated HDNs was measured and no discrimination was made between both types of HF (rEF and pEF) patients^27^. Herein, based on the percentage of H3Cit protein expression, we observed that both HDNs and LDNs from HV and HF are already undergoing NETosis post-isolation. However, the intensity of H3Cit protein expression was much higher on LDNs (up to 11-fold) from all HF patients compared to HDNs, which is in line with the high levels of NETs biomarkers detected in blood from these patients.

When isolated HDNs and LDNs were incubated without agonists, we observed that both neutrophil subtypes from all HF patients were more primed than HV to undergo NETosis. In a previous study, we showed the capacity of CRP to promote NETosis on HDNs from HV and HF patients. When HDNs were stimulated with CRP-depleted serum, NETs formation was abrogated^27^. Herein, we treated both isolated HDNs and LDNs with PMA and CRP and observed that LDNs were more potent to promote NETosis, which was maximal in isolated LDNs from ADHFpEF patients. Interestingly, although CRP (5 mg/L) was less potent than PMA to induce NETs formation in HDNs from all volunteers (HV and HF patients), CRP was equipotent to the powerful pharmacological agonist (PMA) to promote NETs formation in LDNs from ADHFpEF patients. Since plasma levels of CRP in HF patients exceeded the *in vitro* concentration that induced significant NETosis in LDNs and considering that LDNs counts were highest in cases of decompensated HF, our findings strongly suggest that LDNs play a significant role in the observed NETs burden in this context.

We reported that NETs induce adhesion of neutrophils onto hECM, which can be abrogated by DNase I treatment^16^. NETs can also modulate a rapid functional upregulation of neutrophil β_2_-integrin (CD11b/CD18) complex contributing to increased neutrophil adhesion onto hECM^16, 35^. Herein, we observed that isolated HDNs and LDNs from HF patients were more pro-adhesive onto hECM compared to HV, with LDNs from ADHFpEF being the most responsive. In HDNs and LDNs from healthy volunteers only a combination of blocking CD18 Ab and DNase I treatment was efficient to reduce neutrophil adhesion onto hECM, suggesting that these neutrophils were minimally activated. In HF patients, maximal neutrophil adhesion under PMA or CRP treatment occurred in HDNs and LDNs from ADHFpEF patients. Pretreatment either with blocking CD18 Ab or DNase I alone were equipotent to reduce significantly both HDNs and LDNs adhesion, and their combination reduced neutrophil adhesion even below their basal adhesion. These data demonstrate that in ADHFpEF patients, both neutrophil subtypes have a higher cell surface expression of NETs and activated CD11b/CD18 complex, which is in agreement with a recent study reporting that β2 integrin subunit was predominantly overexpressed in HFpEF as opposed to HFrEF patients^36^.

Together, our data demonstrate that not only that there is an increase of LDNs counts in all HF patients, but also that these LDNs correlate with circulating pro-inflammatory cytokines (IL-6, IL-8, CRP) which can promote NETosis, thus, becoming more thrombogenic (increased neutrophil adhesiveness) in presence of elevated NETs-associated biomarkers (MPO-DNA, H3Cit and MPO).

Finally, we observed higher levels of LDNs and NETs-related activity in HFpEF patients, in contrast to HFrEF patients, who typically exhibit poorer clinical prognosis^37^. This finding suggests the existence of distinct characteristics related to inflammation in the mechanisms underlying HFpEF and the resulting myocardial changes^38^. Previous studies have linked neutrophils and NETs-related activity with microvascular dysfunction, a mechanism that plays a crucial role in HFpEF^39^. Consequently, it is conceivable that LDNs may exert differing influences on HF pathogenesis and clinical outcomes according to HF subtype.

### Limitations

In this cross-sectional study, no age-matching was performed in HV. Nonetheless, the changes identified in LDNs among individuals with HF, encompassing various stages of the condition, diminish the likelihood of these alterations being linked to age-related factors. Longitudinal assessment of neutrophils phenotype should be performed to confirm alterations resulting from acute HF decompensation.

### Conclusion

HFpEF patients present higher levels of circulating LDNs and NETs related activities, which are the highest in the context of acute HF decompensation. These LDNs have a higher inflammatory propensity than HDNs as demonstrated by their increased NETosis profile and hECM adhesiveness. Finally, there is a significant increase in systemic NETs biomarkers and inflammatory inducers of NETs release which are all positively correlating with the number of circulating LDNs.

## Abbreviations

ADHF: acute decompensated heart failure
H3Cit: citrullinated histone H3
HDNs: high density neutrophils
LDNs: low density neutrophils
MPO: myeloperoxidase
NETs: neutrophil extracellular traps
pEF: preserved ejection fraction
PMA: phorbol myristate acetate
rEF: reduced ejection fraction
vWF: von Willebrand factor

## Data Availability

The authors declare that all supporting data are available within the article and its online supplementary files.

